# The serological diversity of serum IgG/IgA/IgM against the SARS-CoV-2 nucleoprotein, spike, and receptor-binding domain and neutralizing antibodies in patients with COVID-19 in Japan

**DOI:** 10.1101/2021.06.17.21258858

**Authors:** Yudai Kaneko, Akira Sugiyama, Toshiya Tanaka, Kazushige Fukui, Akashi Taguchi, Aya Nakayama, Kazumasa Koga, Yoshiro Kishi, Wang Daming, Chungen Qian, Fuzhen Xia, Fan He, Liang Zheng, Yi Yu, Youichiro Wada, Yoshiaki Wada, Tatsuhiko Kodama, Takeshi Kawamura

**Affiliations:** Laboratory for Systems Biology and Medicine, Research Centre for Advanced Science and Technology (RCAST), The University of Tokyo, Tokyo, Japan; Medical & Biological Laboratories Co., Ltd, Tokyo, Japan; Isotope Science Centre, The University of Tokyo, Tokyo, Japan; Department of Neurology, Nissan Tamagawa Hospital, Tokyo, Japan; Suzhou Institute of Biomedical Engineering and Technology Chinese Academy of Sciences, Suzhou, China; The Key Laboratory for Biomedical Photonics of MOE at Wuhan National Laboratory for Optoelectronics – Hubei Bioinformatics & Molecular Imaging Key Laboratory, Systems Biology Theme, Department of Biomedical Engineering, College of Life Science and Technology, Huazhong University of Science and Technology, Hubei, China; Reagent R&D Centre, Shenzhen YHLO Biotech Co., Ltd, Guangdong, China

**Keywords:** SARS-CoV-2, COVID-19, antibody, neutralizing antibody, patient

## Abstract

**Objectives:** To compare the temporal changes of IgM, IgG, and IgA antibodies against the SARS-CoV-2 nucleoprotein, S1 subunit, and receptor binding domain and neutralizing antibodies (NAbs) against SARS-CoV-2 in patients with COVID-19.

**Methods:** A total of five patients in Nissan Tamagawa Hospital, Tokyo, Japan confirmed COVID-19 from August 8, 2020 to August 14, 2020 were investigated. Serum samples were acquired multiple times from 0 to 76 days after symptom onset. Using a fully automated CLIA analyzer, we measured the levels of IgG, IgA, and IgM against the SARS-CoV-2 N, S1, and RBD and NAbs against SARS-CoV-2.

**Results:** The levels of IgG antibodies against SARS-CoV-2 structural proteins increased over time in all cases but IgM and IgA levels against SARS-CoV-2 showed different increasing trends among individuals in the early stage. In particular, we observed IgA antibodies increasing before IgG and IgM in 3/5 cases. The NAb levels against SARS-CoV-2 increased and kept above 10 AU/mL more than around 70 days after symptom onset in all cases. Furthermore, in the early stage, NAb levels were more than cut off value in 4/5 COVID-19 patients some of whose antibodies against RBD didn’t exceed 10 AU/mL.

**Conclusions:** Our findings indicate that patients with COVID-19 should be examined for IgG, IgA and IgM antibodies against SARS-CoV-2 structural proteins and NAbs against SARS-CoV-2 in addition to conventional antibody testing methods for SARS-CoV-2 (IgG and IgM kits) to analyze the diversity of patients’ immune mechanisms.

## Introduction

COVID-19 caused by the novel coronavirus severe acute respiratory syndrome coronavirus 2 (SARS-CoV-2) has caused a worldwide pandemic [1]. SARS-CoV-2 is a positive-sense single-stranded RNA virus of the Coronaviridae family that is composed of an envelope (E), membrane (M), nucleoprotein (N), and spike (S) [2]. Coronaviruses entry and infect cells by binding the receptor binding domain (RBD) in the S1 subunit of their trimeric spike proteins to angiotensin-converting enzyme 2 (ACE2) on cell surfaces [3] so a number of neutralizing antibodies (NAbs) against SARS-CoV-2 were reported to target the RBD and block the binding between RBD and ACE2 [4, 5]. As understanding the humoral immunity is important for developing drugs and vaccines for COVID-19, measuring the levels of antibodies against SARS-CoV-2 structural proteins and NAbs against SARS-CoV-2 over time will help identify key factors involved in the immune response. Measurements and monitoring of antibodies (mainly IgG and IgM) against SARS-CoV-2 have already been performed, and some study reported that antibodies are useful diagnostic tools for SARS-CoV-2 infections [6, 7]. However, the longitudinal measurements of antibody isotypes against the SARS-CoV-2 structural proteins and NAbs against SARS-CoV-2 in individuals have not been performed. Therefore, in the present study, we measured chronological changes in the IgG, IgA, and IgM antibodies against the SARS-CoV-2 N, S1, and RBD and NAbs against SARS-CoV-2.

## Methods

### Reagents and instruments

2019-nCoV IgM and IgG chemiluminescence immunoassay (CLIA) kits, detecting SARS-CoV-2 recombinant antigen, were purchased from Shenzhen YHLO Biotech Co., Ltd (Shenzhen, China). Covid-2019 IgG Kit against N antigen (recombinant N protein), Covid-2019 IgM Kit against N antigen (recombinant N protein), Covid-2019 IgA Kit against N antigen (recombinant N protein), Covid-2019 IgG Kit against S1 antigen (recombinant S1 protein), Covid-2019 IgM Kit against S1 antigen (recombinant S1 protein), Covid-2019 IgA Kit against S1 antigen (recombinant S1 protein), Covid-2019 IgG Kit against RBD antigen (recombinant RBD protein), Covid-2019 IgM Kit against RBD antigen (recombinant RBD protein), Covid-2019 IgA Kit against RBD antigen (recombinant RBD protein) and iFlash-2019-nCoV NAb, a one-step competitive immunoassay, were obtained from Shenzhen YHLO Biotech Co., Ltd (Shenzhen, China). A fully automatic CLIA analyzer (iFlash3000) was purchased from Shenzhen YHLO Biotech Co., Ltd. We summarized all kits for immunoassays on Table 1.

**Table 1.**
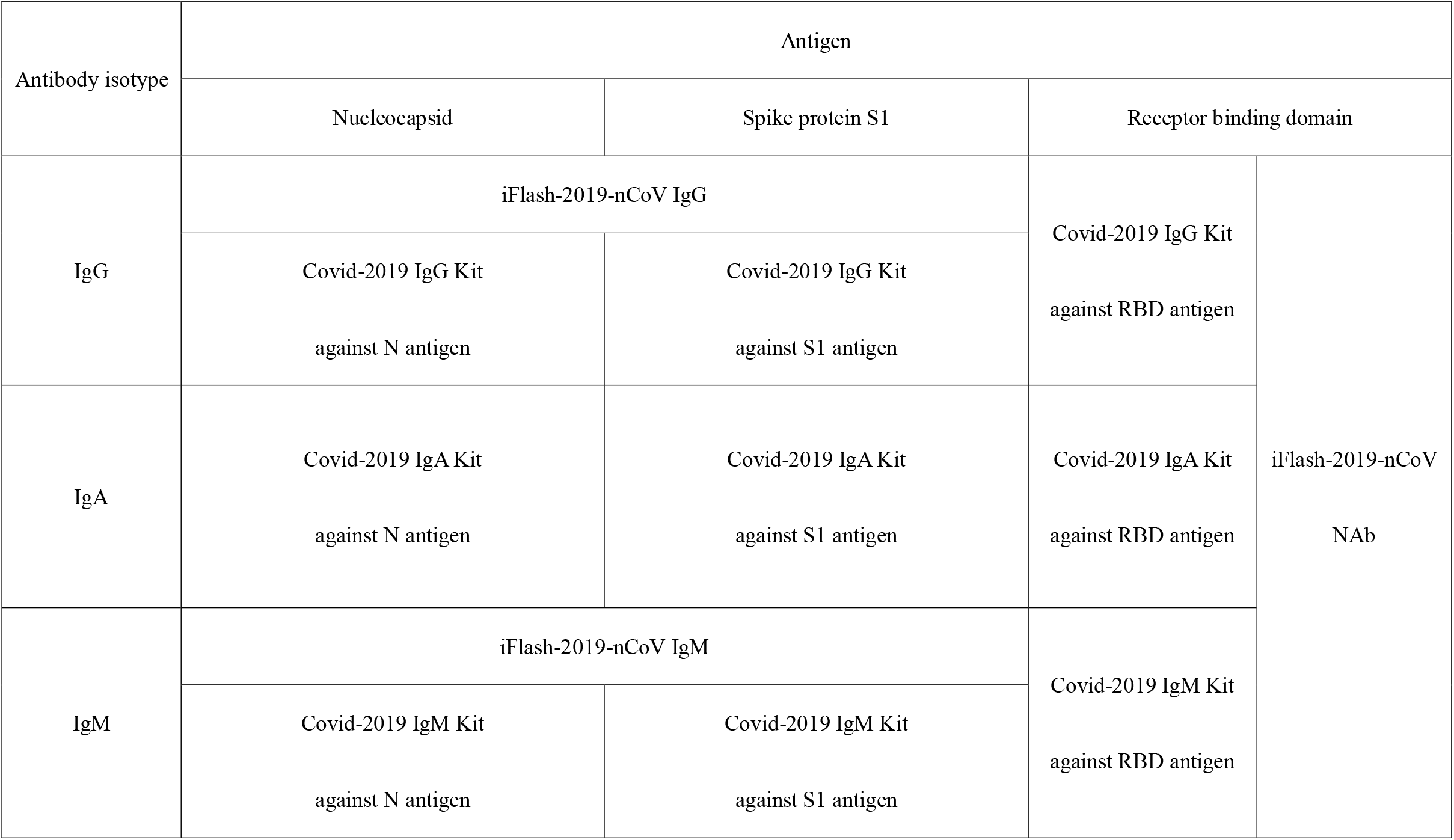
Reagents for detecting antibody isotypes against SARS-CoV-2 structural proteins

### Patients

A total of five patients confirmed COVID-19 in Nissan Tamagawa Hospital from August 8, 2020 to August 14, 2020 were enrolled in this study. At least seven serum samples for each patient were collected from 0 to 76 days after symptom onset. This study was performed at the University of Tokyo and Nissan Tamagawa Hospital approved by their ethics committee (protocol number R2-05 and Tama2020-003).

### Serological test

Levels of IgG, IgA, and IgM against SARS-CoV-2 N, S1, and RBD and neutralizing antibody (NAb) against SARS-CoV-2 were measured using a fully automatic CLIA analyzer (iFlash3000). The antibody concentrations (AU/mL) were calculated based on the relative light units (RLU) obtained by the CLIA analyzer. The cut-off value for indicating a positive test result as used by the manufacturer for all kits was 10 AU/mL.

## Results

The subjects of this study were five men in their 20s to 50s with COVID-19. All infections were confirmed at Nissan Tamagawa Hospital. Table 2 shows the symptoms exhibited by these patients. The most common symptom was fever, with other symptoms including sore throat, cough, dysgeusia, and headache. Patient 2 and Patient 4 exhibited pneumonia. We didn’t find correlation between symptom severity and Real Time PCR Ct values of SARS-CoV-2.

**Table 2.** Symptoms exhibited by the patients in this study

|  |  |  |  | Symptoms |  |  |  |  |  |  |
| --- | --- | --- | --- | --- | --- | --- | --- | --- | --- | --- |
| No. | Age range | Sex | Ct value | Fever | Cough | Sore throat | Dysgeusia | Headache | Diarrhoea | Dyspnea |
| P1 | 20 – 29 | Male | 32.9 | Yes | Yes | Yes | No | No | Yes | No |
| P2 | 40 – 49 | Male | 26.6 | Yes | No | No | No | Yes | No | Yes |
| P3 | 30 – 39 | Male | 22.3 | No | No | No | Yes | No | No | No |
| P4 | 50 – 59 | Male | 18.6 | Yes | No | Yes | No | No | No | Yes |
| P5 | 30 – 39 | Male | 30.5 | Yes | No | No | No | No | No | No |

IgG and IgM levels against SARS-CoV-2 were measured using a fully automatic CLIA analyzer (iFlash3000) (Fig. 1). The IgM seroconversion time was slower than IgG. IgG levels against SARS-CoV-2 increased after symptom onset, exceeding 10 AU/mL as early as 7 days (Fig. 1D) and as late as 17 days after symptom onset (Fig. 1E). In patients with pneumonia, it took a mean of 7 days for IgG antibody levels against SARS-CoV-2 to exceed 10 AU/mL (Fig. 1B, D) while in other patients it took a mean of 16 days (Fig. 1A, C, E). In addition, IgG levels in patients with pneumonia tended to increase over a long period while IgG levels in those with mild symptoms exhibited a decreasing trend starting from day 18. IgM antibody levels against SARS-CoV-2 in only two patients with pneumonia exceeded 10 AU/mL (Fig. 1B, D) while IgM levels against SARS-CoV-2 in other patients were lower than the cut-off value.

**Figure 1.**
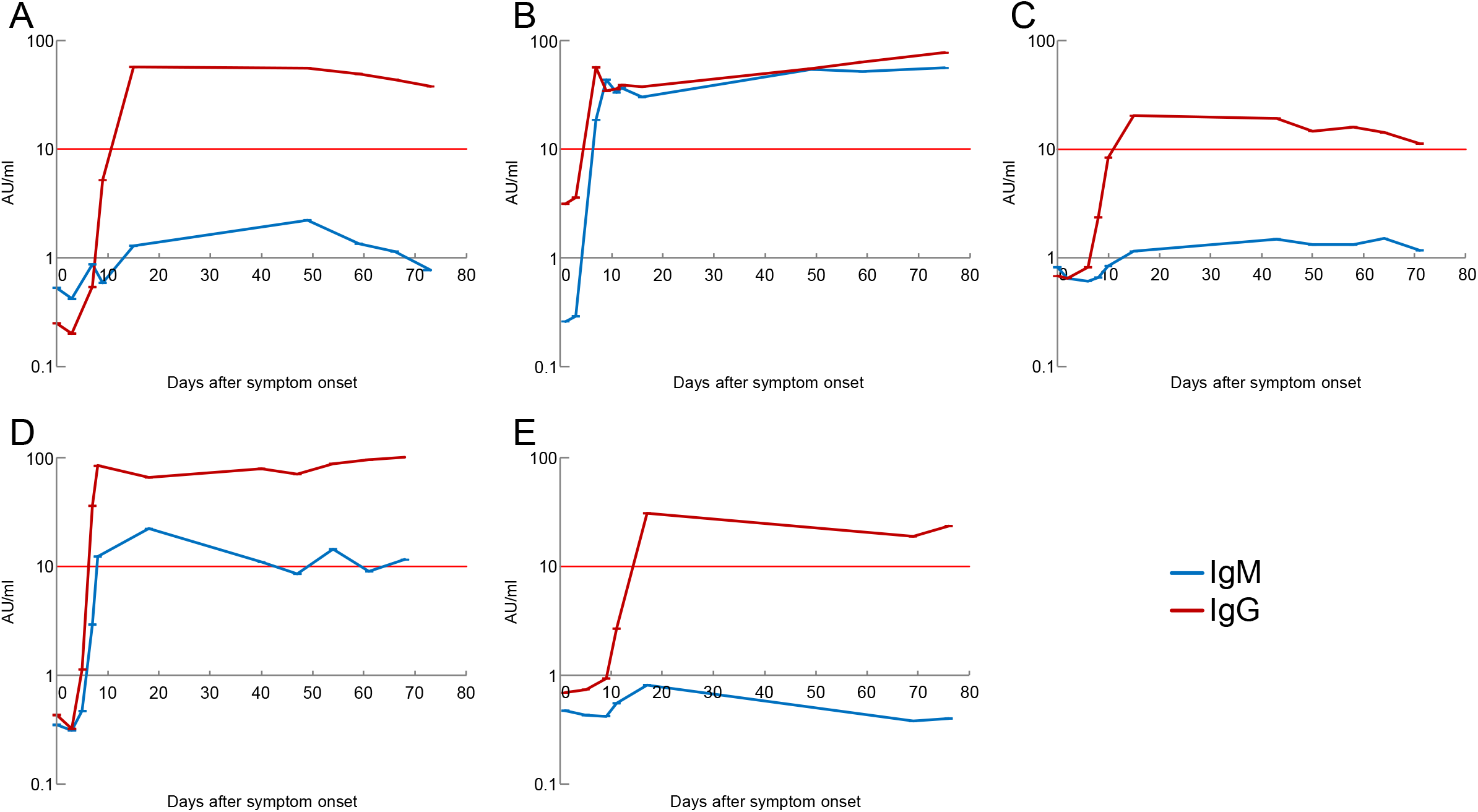
Timeline of IgG/IgM antibodies level against SARS-CoV-2 after symptom onset in patient P1(A), P2(B), P3(C), P4(D) and P5(E).

To evaluate antibodies against SARS-CoV-2 in greater detail, we measured levels of IgG, IgA, and IgM against the N, S1, and RBD and NAbs against SARS-CoV-2 using a fully automatic CLIA analyzer (iFlash 3000) (Fig. 2). IgG against N and S1 increased in the early stage, exceeding 10 AU/mL as early as day 7 (Fig. 2B) and as late as day 17 (Fig. 2E) after symptom onset. In addition, it took 7 to 9 days for IgG against N and S1 to exceed 10 AU/mL in patients with pneumonia (Fig. 2B, D) but it took 15 to 17 days in patients with mild symptoms (Fig. 2A, C, E). In contrast, levels of IgM and IgA against N and S1 exhibited different increasing trends among patients in the early stage. For example, in Patient 5, IgM and IgG levels were low but IgA levels were high on day 11. Furthermore, in Patient 3 and Patient 5 who only exhibited one symptom, levels of IgA against N were already slightly elevated on day 1 after symptom onset. In later stage, IgG levels remained flat followed by rise in all samples while the levels of IgA fell gradually and below the cut-off value around day 70 in 4/5 patients. The changes in levels of IgG, IgA, and IgM against RBD were similar to those of IgG, IgA, and IgM against S1 from day 20 after COVID-19 symptom onset although the increasing trends of antibody levels against S1 weren’t different from those of the antibody levels against RBD in early stage.

**Figure 2.**
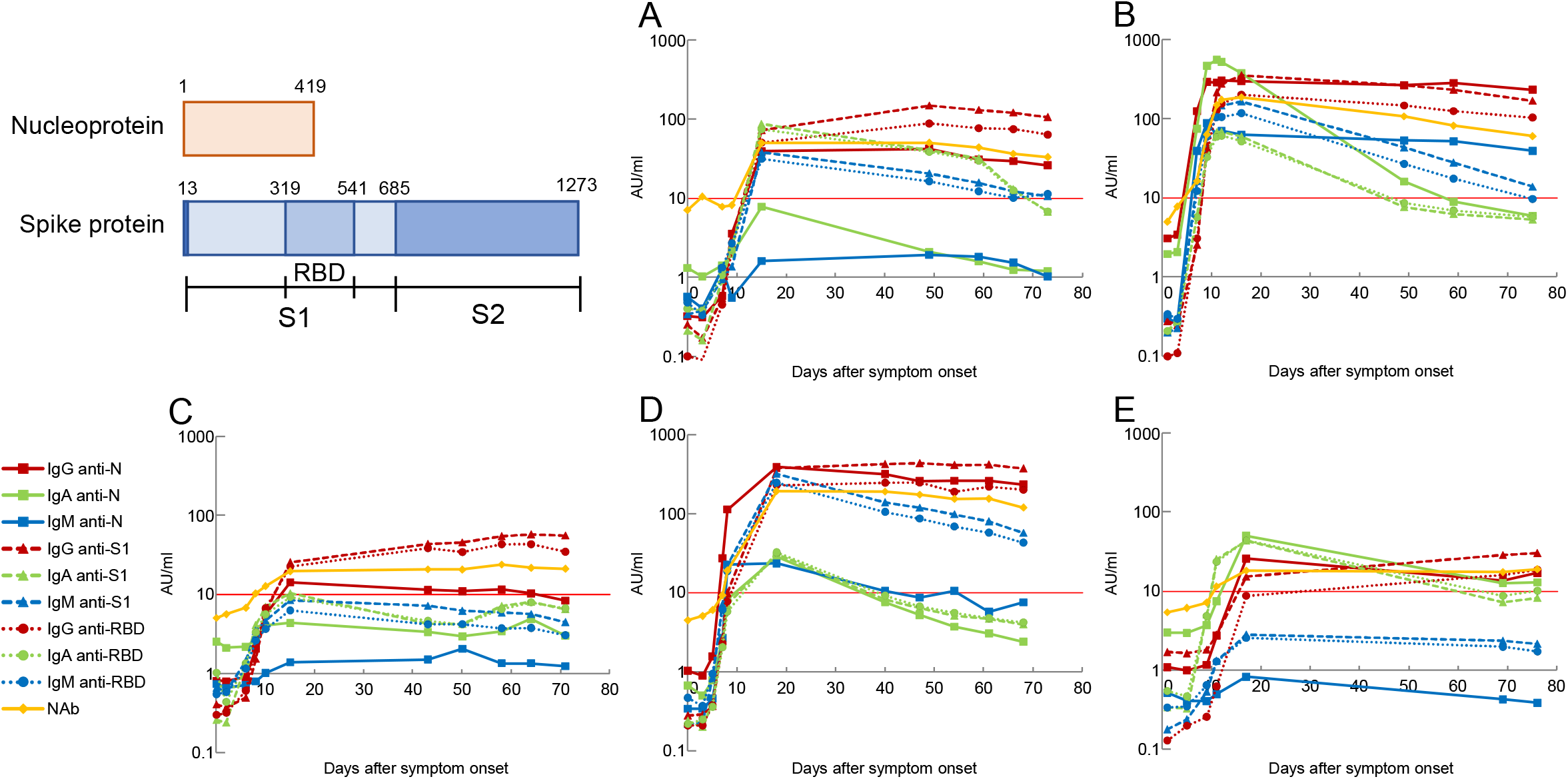
Timeline of IgG/IgA/IgM antibody levels against N/S1/RBD antigens of SARS-CoV-2 and neutralizing antibody against SARS-CoV-2 after symptom onset in patient P1(A), P2(B), P3(C), P4(D) and P5(E).

NAbs against SARS-CoV-2 tended to increase, exceeding 10 AU/mL less than 15 days after symptom onset (Fig. 2). The peak levels of NAbs against SARS-CoV-2 in patients with pneumonia were much higher than NAbs levels in those with mild symptoms. NAb levels stayed above 10 AU/mL more than around 70 days after symptom onset while the levels of IgA and IgM against RBD dropped below 10 AU/mL around 70 days after symptom onset in 3/5 patients (Patient 2, Patient 3, and Patient 5). Furthermore, in the early stage, one or no antibody isotype against RBD exceeded 10 AU/mL in 4/5 COVID-19 patients whose NAb levels were more than 10 AU/mL. For example, in Patient 2 and Patient 4, only IgM levels against RBD were more than the cut-off value when the levels of Nabs exceeded 10 AU/mL on day 7 and 8 after symptom onset, respectively. In Patient 3, the levels of any antibody isotypes against RBD were less than 10AU/ml when NAb levels over the cut-off value from Day 9 to Day 11 after symptom onset.

To analyze antibodies against SARS-CoV-2 in COVID-19 patients, we compared levels of IgG, IgA, and IgM against the N, S1, and RBD and NAbs against SARS-CoV-2. We observed the higher correlation between the levels of antibodies detected by using 2019-nCoV IgG and IgM CLIA kits and anti-N antibodies than anti-S and anti-RBD antibodies (anti-N IgG: R^2=0.8804, anti-S1 IgG: R^2=0.7919, anti-RBD IgG: R^2=0.8196, anti-N IgM: R^2=0.9727, anti-S1 IgM: R^2=0.6832, anti-RBD IgM: R^2=0.6949) (Fig3A, B). Furthermore, there are very high correlations between IgG, IgA, and IgM antibodies against S1 and RBD (IgG: R^2=0.9612, IgA: R^2=0.9794, IgM: R^2=0.9808) (Fig. 3C). The NAb levels were correlated most strongly with IgG levels of the antibody isotypes (anti-N IgG: R^2=0.8447, anti-S1 IgG: R^2=0.9127 and anti-RBD IgG: R^2=0.9251, anti-N IgA: R^2=0.2226, anti-S1 IgA: R^2=0.2087, anti-RBD IgA: R^2=0.2202, anti-N IgM: R^2=0.3115, anti-S1 IgM: R^2=0.7947 and anti-RBD IgM: R^2=0.7657) (Fig. 4).

**Figure 3.**
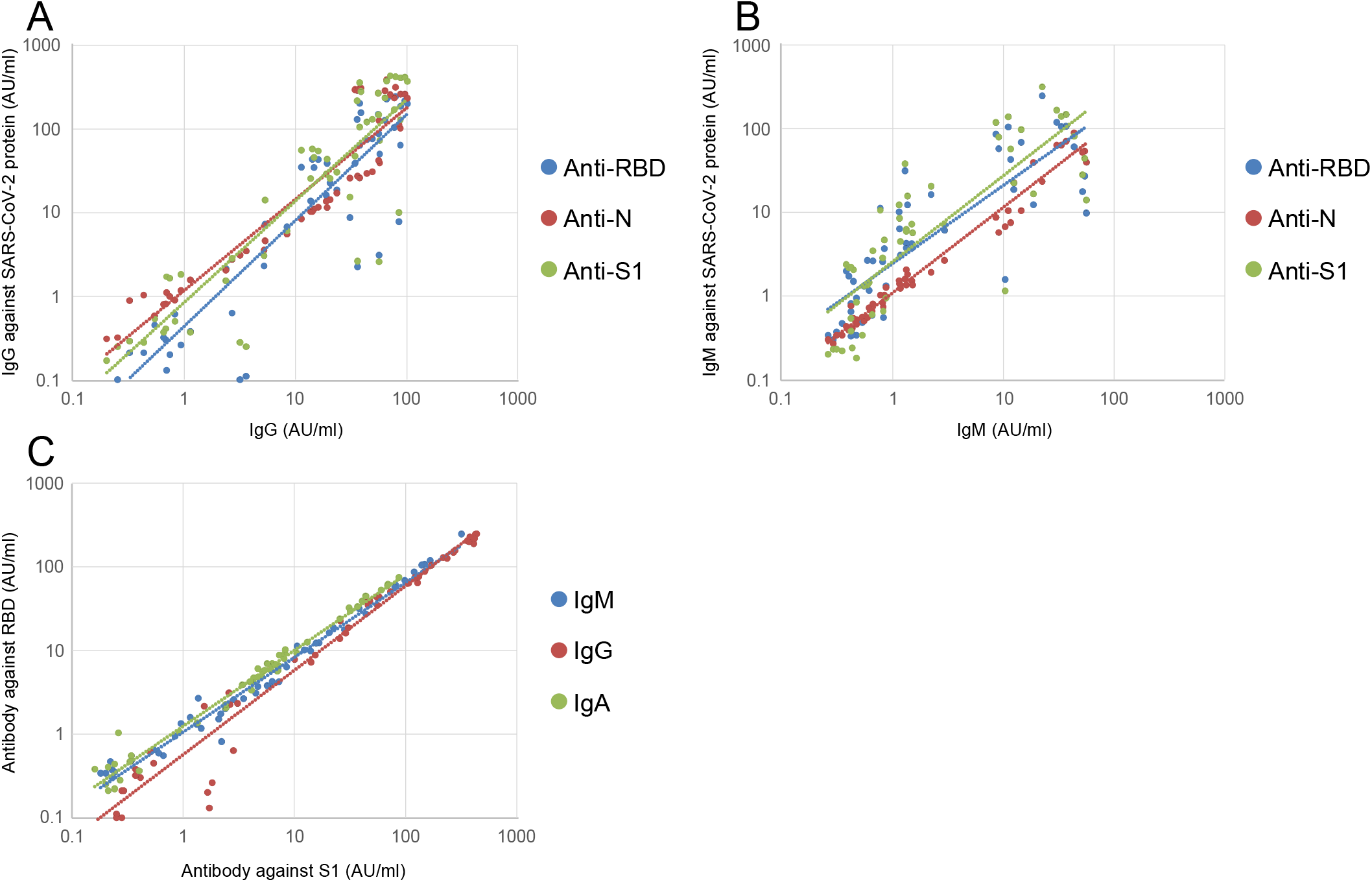
Correlations between IgG(A) /IgM(B) levels and the IgG/IgM against N, S1 and RBD. Correlations between IgG, IgA and IgM levels against S1 and those against RBD (C).

**Figure 4.**
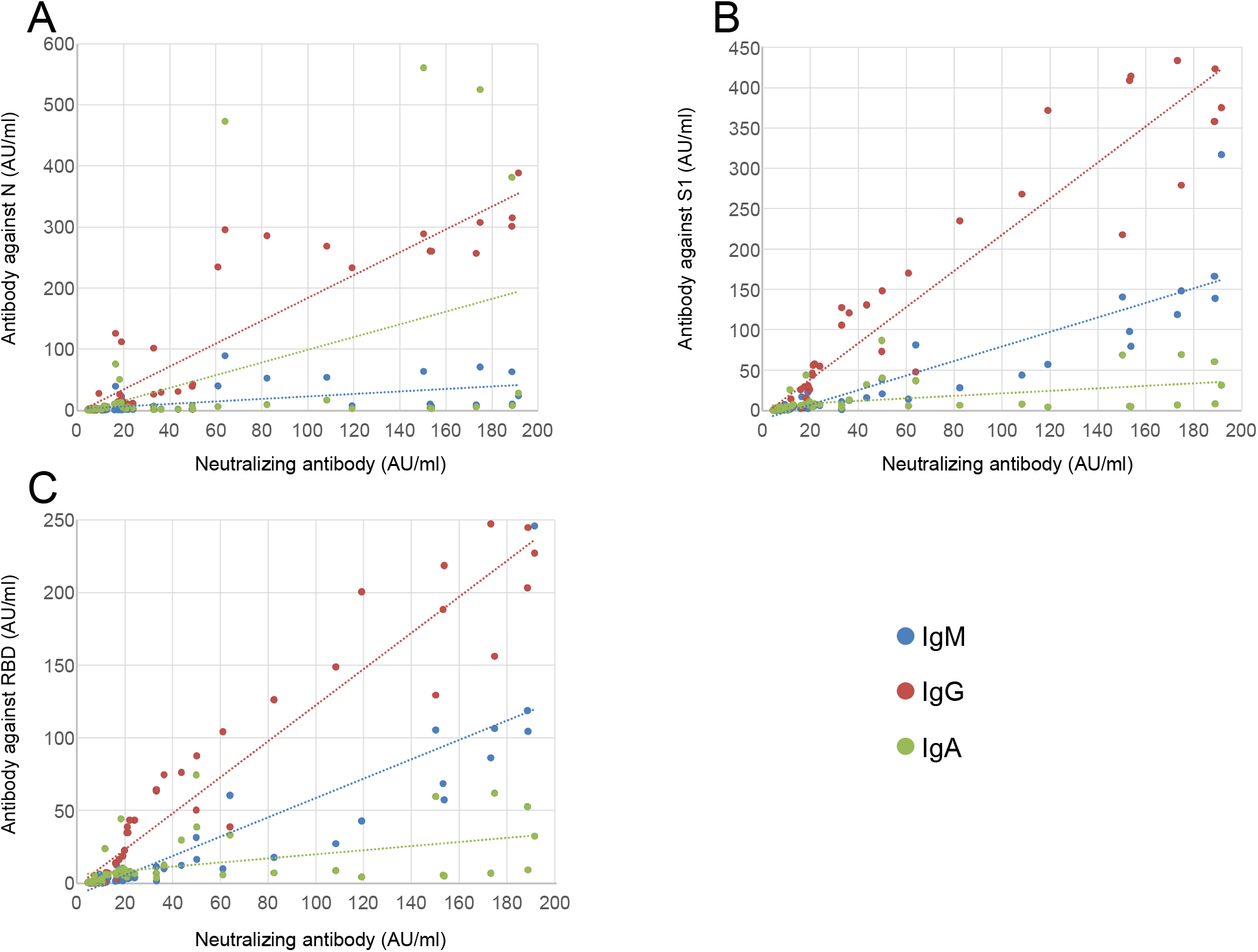
Correlations between NAb levels and antibody levels against SARS-CoV-2 N(A), S1(B) and RBD(C).

## Discussion

In this study, we examined the levels of IgG, IgA, and IgM antibodies against SARS-CoV-2 N, S1, and RBD and NAbs against SARS-CoV-2 in patients’ sera with COVID-19 using a fully automated CLIA analyzer. Our results demonstrated the serological diversity of serum IgG, IgA, and IgM antibodies against the SARS-CoV-2 N, S1 and RBD and NAbs against SARS-CoV-2 in patients with COVID-19 and the necessity of combinational measurements of the antibody isotype levels against each structural protein of SARS-CoV-2 and NAb levels.

Using 2019-nCoV IgG kits and 2019-nCoV IgM kits, the chronological change of IgG antibody in patients with COVID-19 found in this study is in line with other recent publications. Several reports showed that IgG levels were higher in the later stages after symptom onset in severe cases than those in mild cases and that patients who die from COVID-19 exhibit higher IgM levels [8,9]. Our results show that IgG antibody levels against SARS-CoV-2 in the later stage and IgM antibody levels against SARS-CoV-2 could be useful for diagnosing the severity of the disease. Our results showed that seroconversion time of IgM was slower than that of IgG in COVID-19 patients’ sera after symptom onset. A recent study also reported that IgM seroconversion was later than that of IgG in some cases [10], not being consistent with the typical model that IgM seroconversion time is faster than IgG in the early stage after viral infections. These data suggest that individuals who were not confirmed SARS-CoV-2 infection may already have antibodies possessing cross-reactivity with SARS-CoV-2 in some cases, being infected by a member of the coronavirus family.

The IgG and IgM levels were correlated strongly with the antibody levels against SARS-CoV-2 N proteins. In particular, we observed the high correlation between the levels of antibodies measured by using 2019-nCoV IgG and IgM CLIA kits and anti-N antibodies at low antibody levels. According to the manufacturer, 2019-nCoV IgG and IgM CLIA kits detect antibodies against the SARS-CoV-2 recombinant antigen. We speculate that magnetic microparticles are mainly coated with N antigens. In order to evaluate not only anti-N antibodies but also anti-S1 antibodies accurately, we need use the kit detecting the levels of antibodies against SARS-CoV-2 structural protein as well as conventional testing kit (2019-nCoV IgG/IgM kit).

Using Covid-2019 IgG, IgA, and IgM kit against N, S1, and RBD antigen, levels of IgG against SARS-CoV-2 increased after symptom onset in all patients with COVID-19 but some cases exhibited different seroconversion times such as IgA levels rising before IgG and IgM. A recent study reported that IgA levels in serum increased soon after symptom onset with mild symptoms while that a case with the severity of symptoms showed a delayed but very strong IgA response against SARS-CoV-2 [11]. Furthermore, measurement of serum IgA, in addition to IgM and IgG, improved diagnostic accuracy for SARS-CoV-2 infections [12]. We also observed higher levels of NAbs in patients with severe symptom than NAb levels in patients with mild symptom, using 2019-nCoV NAb kits. The NAb levels were reported to increase after SARS-CoV-2 infection in most individuals [13] and be associated with clinical disease severity [14, 15], confirming the results of our studies. IgA and NAb levels against SARS-CoV-2 could be biomarker for COVID-19 severity.

There is the highest correlation between NAb and IgG levels against RBD of antibody isotypes in this study. Furthermore, IgG levels against RBD are maintained in later stage, exhibiting similarly changes of NAb levels, while the ratios of NAb levels to IgA and IgM levels against RBD continuously increased starting on day 18 after COVID-19 symptom onset in all cases. Our data suggested that isotype switching to IgG could take place and serum IgG mainly could have neutralizing activity against SARS-CoV-2 in the later stage, being consistent with a recent study [16].

The NAb kit offer us some advantages by evaluating all antibodies having neutralization activity in the COVID-19 patients. First, the NAb kit detects ACE2 competitively binding to RBD-coated particles with antibodies against SARS-CoV-2 structural proteins while typical antibody kit against structural proteins detect each isotype, not reflecting total NAb levels. In fact, we observed the levels of NAbs keeping above the cut off value more than around 70 days after symptom onset in all COVID-19 patients some of whose antibody levels against RBD lower than the cut-off value. Furthermore, some of antibody isotypes against RBD fall below 10AU/ml in early stage while NAb levels exceed the cut-off value in 4/5 patients. Second, the levels of IgG, IgA, and IgM against structural proteins weren’t detected correctly, biased by different affinities and avidity of antibody isotypes. The NAb kit could be as useful as IgG, IgA, and IgM kit against S1 and RBD for the accurate measurement of antibodies having neutralizing activity in patients’ sera with COVID-19.

However, there are some limitations in this study. First, the number of patients is the relatively small this study. It is necessary for us to measure antibody levels in many COVID-19 patients’ sera for a more accurate statistical significance regarding the correlations between antibody isotypes and the severity of symptoms. Second, we didn’t detect neutralizing activity of each isotype against SARS-CoV-2 by the methods considering the bias of different affinities of antibody isotypes as explained above. Furthermore, each antibody isotype affects the total NAb levels at different times after infection because of different seroconversion. We observed IgA levels against S1 and RBD declining followed by rise and below 10AU/mL around day 70 in all cases and a recent study also reported that IgA levels in serum decreased notably 1 month after symptom onset, being more potent than IgG in neutralizing activity for SARS-CoV-2 [17]. We need assess neutralizing activity of IgG, IgA, and IgM against SARS-CoV-2, using the assay which can measure each isotype for understanding the role of these isotype against SARS-CoV-2. Third, we detected NAbs against only RBD of SARS-CoV-2 in serum samples. Antibodies against S1 region except for RBD of SARS-CoV-2 were reported to have neutralizing activity [18, 19]. We should detect NAbs by using S1-coated particles, including RBD region for analyzing neutralization antibody. Last, we found the persistence of NAbs in patients with COVID-19 around 70 days after symptom onset but we haven’t measured the neutralization activity of antibody for a longer time yet. Some longitudinal studies have reported that neutralization activity against SARS-CoV-2 significantly declined as early as 6 weeks and that persisted as late as 5 months after symptom onset [20, 21, 22]. Furthermore, the binding surface in SARS-CoV-2 RBD to ACE2 is reported to be less antigenic than that of other S regions [23] so the antibody levels against RBD could go down earlier than antibody levels against S1, which may affect chronological changes in NAb levels. Further longitudinal analysis of COVID-19 patients is needed to understand immune memory after SARS-CoV-2 infection and vaccine.

## Conclusion

To further elucidate humoral immunity and other aspects of the immune mechanism of COVID-19, we must combine measurements, such as levels of IgG, IgA, and IgM against SARS-CoV-2 N, S1, and RBD and Nabs against SARS-CoV-2, in addition to using conventional testing methods (2019-nCoV IgG/IgM kit).

## Data Availability

The data that support the findings of this study are available from the corresponding author, T. K., upon reasonable request.

## Transparency declaration

The authors declared the following potential conflicts of interest with respect to the research, authorship, and/or publication of this article: 2019-nCoV IgM and IgG chemiluminescence immunoassay (CLIA) kits, Covid-2019 IgG/IgA/IgM Kit against N/S1/RBD antigen, and iFlash-2019-nCoV NAb were sponsored by Medical & Biological Laboratories Co., Ltd. and Shenzhen YHLO Biotech Co., Ltd.

## Funding

This study was supported by grants from the Peace Winds Japan, Kowa Co., the Research Center for Advanced Science and Technology, and Kodama laboratory in the University of Tokyo.

## Acknowledgements

We would like to thank all the healthcare workers involved in treating COVID-19 patients at Nissan Tamagawa Hospital.

## Contribution

Yudai Kaneko performed data curation and analysis and drafted the manuscript. Kazushige Fukui, Akashi Taguchi, and Aya Nakayama performed data. Akira Sugiyama, Toshiya Tanaka, Youichiro Wada, Yoshiaki Wada, Tatsuhiko Kodama, and Takeshi Kawamura administrated experiments in the project and critically reviewed the manuscript. Kazumasa Koga collected samples. Yoshiro Kishi critically reviewed the manuscript. Chungen Qian, Fuzhen Xia, Fan He, Liang Zheng, and Yi Yu developed and provided the IgG, IgA, and IgM kits for SARS-CoV-2 N, S1, and RBD and the NAb kit against SARS-CoV-2. Wang Daming developed and provided the NAb kit against SARS-CoV-2.

## Notes

### Author Declarations

This study was performed at the University of Tokyo and Nissan Tamagawa Hospital approved by their ethics committee (protocol number R2-05 and Tama2020-003).

## References

[1] World Health Organization. (2020) Coronavirus disease (COVID-19) dashboard. Available from: https://covid19.who.int/ Accessed 22nd August 2020

[2] Weiss SR, Leibowitz JL. Coronavirus pathogenesis. Adv Virus Res. 2011;81:85–164, 10.1016/B978-0-12-385885-6.00009-2.

[3] Hoffmann M, Kleine-Weber H, Schroeder S, Krüger N, Herrler T, Erichsen S et al. SARS-CoV-2 Cell Entry Depends on ACE2 and TMPRSS2 and Is Blocked by a Clinically Proven Protease Inhibitor. Cell. 181(2):271-280.e8, 10.1016/j.cell.2020.02.052. (Epub ahead of print)

[4] Shi R, Shan C, Duan X, Chen Z, Liu P, Song J et al. A human neutralizing antibody targets the receptor-binding site of SARS-CoV-2. Nature. 2020;584(7819):120–124. doi: 10.1038/s41586-020-2381-y. (Epub ahead of print)

[5] Cao Y, Su B, Guo X, Sun W, Deng Y, Bao L et al. Potent Neutralizing Antibodies against SARS-CoV-2 Identified by High-Throughput Single-Cell Sequencing of Convalescent Patients’ B Cells. Cell. 2020;182(1):73-84.e16. doi: 10.1016/j.cell.2020.05.025. (Epub ahead of print)

[6] Infantino M, Grossi V, Lari B, Bambi R, Perri A, Manneschi M et al. Diagnostic accuracy of an automated CLIA for anti-SARS-CoV-2 IgM and IgG antibodies: an Italian experience. J Med Virol. 2020; 92(9):1671-1675. 10.1002/jmv.25932. (Epub ahead of print)

[7] Jin Y, Wang M, Zuo Z, Fan C, Ye F, Cai Z et al. Diagnostic value and dynamic variance of serum antibody in coronavirus disease. Int J Infect Dis IJID Off Publ Int Soc Infect Dis 2019;94:49-52. 10.1016/j.ijid.2020.03.065. (Epub ahead of print)

[8] Zhang B, Zhou X, Zhu C, Song Y, Feng F, Qiu Y et al. Immune phenotyping based on the neutrophil-to-lymphocyte ratio and IgG level predicts disease severity and outcome for patients with COVID-19. Front Mol Biosci 2020;7:157, 10.3389/fmolb.2020.00157. (Epub ahead of print)

[9] Wang Z, Li H, Li J, Yang C, Guo X, Hu Z et al. Elevated serum IgM levels indicate poor outcome in patients with coronavirus disease 2019 pneumonia: a retrospective case-control study. medRxiv. 2020; 10.1101/2020.03.22.20041285 (Epub ahead of print)

[10] Long QX, Deng HJ, Chen J, Hu JL, Liu BZ, Liao P et al. Antibody responses to SARS-CoV-2 in COVID-19 patients: the perspective application of serological tests in clinical practice. medRxiv. 2020. 10.1101/2020.03.18.20038018

[11] Dahlke C, Heidepriem J, Kobbe R, Santer R, Koch T, Fathi A et al. Distinct early IgA profile may determine severity of COVID-19 symptoms: an immunological case series. medRxiv. 2020. 10.1101/2020.04.14.20059733

[12] Ma H, Zeng W, He H, Zhao D, Yang Y, Jiang D et al. COVID-19 diagnosis and study of serum SARS-CoV-2 specific IgA, IgM and IgG by chemiluminescence immunoanalysis. medRxiv. 2020; 10.1101/2020.04.17.20064907

[13] Lau EHY, Tsang OTY, Hui DSC, Kwan MYW, Chan WH, Chiu SS et al. Neutralizing antibody titres in SARS-CoV-2 infections. Nat Commun. 2021;12(1):63. 10.1038/s41467-020-20247-4.

[14] Jeewandara C, Jayathilaka D, Gomes L, Wijewickrama A, Narangoda E, Idampitiya D et al. SARS-CoV-2 neutralizing antibodies in patients with varying severity of acute COVID-19 illness. Sci Rep. 2021;11(1):2062. 10.1038/s41598-021-81629-2.

[15] Legros V, Denolly S, Vogrig M, Boson B, Siret E, Rigaill J et al. A longitudinal study of SARS-CoV-2-infected patients reveals a high correlation between neutralizing antibodies and COVID-19 severity. Cell Mol Immunol. 2021 1–10. 10.1038/s41423-020-00588-2. (Epub ahead of print)

[16] Isho B, Abe KT, Zuo M, Jamal AJ, Rathod B, Wang JH et al., Persistence of serum and saliva antibody responses to SARS-CoV-2 spike antigens in COVID-19 patients. Sci Immunol. 2020;5(52):eabe5511. doi: 10.1126/sciimmunol.abe5511.

[17] Sterlin D, Mathian A, Miyara M, Mohr A, Anna F, Claër L et al. IgA dominates the early neutralizing antibody response to SARS-CoV-2. Sci Transl Med. 20213(577):eabd2223. 10.1126/scitranslmed.abd2223. (Epub ahead of print)

[18] Wan J, Xing S, Ding L, Wang Y, Gu C, Wu Y et al, Human-IgG-Neutralizing Monoclonal Antibodies Block the SARS-CoV-2 Infection. Cell Rep. 2020;32(3):107918. doi: 10.1016/j.celrep.2020.107918. (Epub ahead of print)

[19] Chi X, Yan R, Zhang J, Zhang G, Zhang Y, Hao M et al. A neutralizing human antibody binds to the N-terminal domain of the Spike protein of SARS-CoV-2. Science. 2020;369(6504):650–655. 10.1126/science.abc6952. (Epub ahead of print)

[20] Prévost J, Gasser R, Beaudoin-Bussières G, Richard J, Duerr R, Laumaea A et al. Cross-sectional evaluation of humoral responses against SARS-CoV-2 spike. Cell Rep Med 2020;1(7):100126, 10.1016/j.xcrm.2020.100126. (Epub ahead of print)

[21] Wajnberg A, Amanat F, Firpo A, Altman DR, Bailey MJ, Mansour M et al. Robust neutralizing antibodies to SARS-CoV-2 infection persist for months. Science. 2020;370(6521):1227–1230. 10.1126/science.abd7728. (Epub ahead of print)

[22] Jeewandara C, Jayathilaka D, Gomes L, Wijewickrama A, Narangoda E, Idampitiya D et al. SARS-CoV-2 neutralizing antibodies in patients with varying severity of acute COVID-19 illness. Sci Rep. 2021;11(1):2062. 10.1038/s41598-021-81629-2.

[23] Hattori T, Koide A, Panchenko T, Romero LA, Teng KW, Tada T et al. The ACE2-binding interface of SARS-CoV-2 Spike inherently deflects immune recognition. bioRxiv 2020; 10.1101/2020.11.03.365270. (Epub ahead of print)

